# Poking COVID-19: insights on genomic constraints among immune-related genes between Qatari and Italian populations

**DOI:** 10.1101/2021.10.04.21264507

**Authors:** Hamdi Mbarek, Massimiliano Cocca, Yasser Al Sarraj, Chadi Saad, Massimo Mezzavilla, Wadha AlMuftah, Dario Cocciadiferro, Antonio Novelli, Isabella Quinti, Azza AlTawashi, Salvino Salvaggio, Asma AlThani, Giuseppe Novelli, Said Ismail

## Abstract

Host genomic information, specifically genomic variations, may characterize susceptibility to disease and identify people with a higher risk of harm, leading to better targeting of care and vaccination. Italy was the epicentre for the spread of COVID-19 in Europe, the first country to go into a national lockdown and has one of the highest COVID-19 associated mortality rates. Qatar, on the other hand has a very low mortality rate. In this study, we compared whole-genome sequencing data of 14398 adults and Qatari-national to 925 Italian individuals. We also included in the comparison whole-exome sequence data from 189 Italian laboratory confirmed COVID-19 cases. We focused our study on a curated list of 3619 candidate genes involved in innate immunity and host-pathogen interaction. Two population-gene metric scores, the Delta Singleton-Cohort variant score (DSC) and Sum Singleton-Cohort variant score (SSC), were applied to estimate the presence of selective constraints in the Qatari population and in the Italian cohorts. Results based on DSC SSC metrics demonstrated a different selective pressure on three genes (MUC5AC, ABCA7, FLNA) between Qatari and Italian populations. This study highlighted the genetic differences between Qatari and Italian populations and identified a subset of genes involved in innate immunity and host-pathogen interaction.

## Introduction

COVID-19 continues to spread worldwide, with over four million deaths to date and rising. However, this global spread is coupled with stark anomalies in morbidity and mortality. These differences can be seen not only between different populations but also within the same population[1–4].While most of these differences can be attributed to sociodemographic and clinical factors, this is also a unique opportunity to assess associations with host genomes. Host genomic information, specifically genomic variations, may characterize susceptibility to disease and identify people with a higher risk of harm, leading to better targeting of care and vaccination[5–7]. In addition, characterizing these host factors may help identifying and development of adapted drugs and vaccines[8–10]. The scientific community came together with several efforts to investigate how the genomic variation in the host affects disease susceptibility and progress[11, 12]. So far, these large consortia efforts have led to the identification of over 20 loci associated with susceptibility or severity of the disease[13].

Italy was the epicentre for the spread of COVID-19 in Europe and the first country to go into a national lockdown. It had one of the highest COVID-19 associated mortality rates in Europe[4]. At the time of writing, almost five million cases have been confirmed, with a death toll of more than 130 thousand people. Qatar, on the other hand, despite having one of the highest worldwide numbers of laboratory-confirmed cases (36,729 cases per million, by July 2020), has a very low mortality rate (infection fatality rate = 0.91 per 10,000 persons, by July 2020, per WHO COVID-19 mortality classification)[14]. Studies have even suggested that some communities in Qatar have reached herd immunity for SARS-CoV-2 at a proportion of infection of 65-70% [15]. With the development of the first generation of RNA based vaccines[16, 17], along with the more standard adenovirus-based solutions[18, 19], the end of the pandemic seems to be in sight, though we are aware that this is just the beginning of a more bearable coexistence with the virus.

In this study, we focused on genes involved in the immune response, combining them with a dataset of 1500 proteins mostly involved in COVID-19 disease[20] and a subset of genes already identified as linked to COVID-19 susceptibility and progression[7]. We applied, on this set of genes, a prioritization method based on ultra-rare and population-specific variants. With this approach, we aim to identify a group of genes showing different signs of selective pressure in our study cohorts. Our hypothesis is that those genes can provide information to understand the pandemic progression and maybe help towards therapy.

## Materials and Methods

### Population description

#### The Qatari Cohort

the Qatar Genome Program (QGP)[21] is a population-based project launched by the Qatar Foundation to generate a large-scale whole-genome sequence (WGS) dataset, in combination with comprehensive phenotypic information collected by the Qatar Biobank (QBB)[22]. All subjects included in the analysis were of Qatari Middle Eastern Arabian ancestry (Razali et al. in press). In this study, we use a cohort of 14,398 individuals with an average coverage of 30x. Data preprocessing and downstream quality control analyses for WGS data were conducted as recommended by the Covid19 Host Genetics Initiative study protocol[23].

#### Italian Isolated cohorts

Three Italian cohorts belonging to the Italian network of Genetic Isolates (INGI) were involved in this study due to the availability of whole genome-sequence data. The selected populations localized in three different geographical areas of Italy: North-West (Val Borbera-VBI), North-East (Friuli Venezia Giulia-FVG) and South-East (Carlantino-CAR); In each cohort, a wide range of phenotypic data was collected for each participant (anthropometric traits, blood tests, sensory impairment, taste and food preferences, extensive personal and familial anamnesis). A total of 925 samples with low coverage (4x to 10x) WGS data were selected for the analyses[24].

#### Italian COVID-19 positive samples

a cohort of 189 samples which tested positive for the SARS-CoV-2 infection and collected at the Bambino Gesu’ hospital in Rome was included in the study to provide information on the pattern of genetic variation in a group of selected genes in an outbred Italian cohort. Whole Exome Sequencing data was generated by the University of Tor Vergata. The samples are clustered in three groups, based on the disease severity: severe, extremely severe and asymptomatic[25].

All data analyzed was aligned to the reference genome’s GRCh38 release, and functional annotations were obtained using the Ensembl VEP tool[26].

### Principal component analysis

To highlight the study cohorts’ population structure level, we performed a principal component analysis (PCA) using KING software[27]. Plink v1.9 software[28] was used to convert data from vcf to plink binary format. QGP and each INGI cohort results were projected into the 1000Genomes Project data[29]. To highlight the peculiar ancestry structure of the Qatari population, we also performed an ancestry inference analysis using the software KING.

### Genes selection and prioritization analyses

#### Literature curation process, GEL panel expert and IVA

The candidate gene generation process, the initial candidate gene list ranking and curation are conveyed on the recent literature review to extract a list of genes involved in innate immunity and host-pathogen interaction. The primary gene list is curated according to the knowledge-literature base by the Ingenuity® Variant Analysis™ software from QIAGEN[30] and the viral gene panel expert from Genomics England (GEL)[31]. This list includes a total of 3617 genes (Table S1).

Candidate genes were annotated with the most common gene-ranking metrics using the loss of function intolerance score (pLI)[32] and the Residual Variation Intolerance Score (RVIS)[33]. In addition, we selected a list of 25 genes (Table S2) that were recently associated with COVID-19 susceptibility and severity[7] and overlapped with the primary list, extracting a subset of genes that underwent further analyses.

#### Population-based Gene constraints

Two population-based gene metric scores, the Delta Singleton-Cohort variant score (DSC–accounting for the difference in singletons between coding and non-coding regions) and Sum Singleton-Cohort variant score (SSC– accounting for the sum of singletons variants in the coding and non-coding regions), were adopted to estimate the presence of specific pressures selection in the Qatari population as well as the Italian Isolated cohorts[34]. Only variants with a QUAL value above 30 were used in the calculation to limit the inclusion of genotyping error for variants with allele count (AC) equal to 1. Only scores calculated on canonical transcripts were selected. Only genes with scores values lower or equal to −2 and greater or equal to 2 were retained in each population. These values represent the significant threshold that allows us to discriminate between a gene under constraint (DSC or SSC score ¡= −2) or under relaxation (DSC or SSC score ¿= 2). Since we aim to compare two populations with different structures and high levels of inbreeding, we also calculated the same set of scores for the closest ancestry populations of each study cohort. We used data from the gnomAD v3.1[35] call set, including 1000Genomes project samples and extracted information on the EUR, AFR and SAS superpopulations subset. The EUR subset was used as reference for the Italian samples[36], while the AFR and SAS subsets for the Qatari cohort [Razali et al, 2021, in press]. We defined two different levels of comparison: at the ancestry level, in which we selected all genes showing concordant selective signals between our study cohorts and their closest ancestry population, and at the population level, in which we selected only genes showing different behaviour between our target population and the reference. Since we were dealing with three Italian populations, comparing only with one reference, we selected genes that satisfied our criteria in at least one of the three target Italian populations.

On the other hand, we used two different references for the Qatari target population, so we selected all genes that meet our criteria in at least one reference population. Finally, we proceeded with the comparisons between our target populations, performing three sets of comparisons: population-specific, population-specific versus ancestry related and ancestry related comparisons (Table S3). Each comparison was performed separately for SSC and DSC scores. Finally, we generated a list of genes overlapping between SSC and DSC comparisons to select those genes that consistently showed opposite behaviour in terms of selection or relaxation in our target populations.

We used the Fisher’s test to compare DSC and SSC scores distributions between study cohorts, and reference populations. We performed a Shapiro-Wilk test to assess the normality of the score distribution in each cohort and an enrichment test to assess whether there was an enrichment in relaxed or constrained genes in our target populations versus the selected reference populations.

#### WES Covid-19 cohorts

Using Whole Exome Sequence data from a cohort of Covid-19 positive samples (n=189), we calculated singletons count and singletons density in the coding regions of the genes belonging to the shortlist generated, adjusting by sample size, and compared using Fisher’s test against data from the other target and reference populations. In this cohort, each sample was characterized by a disease severity code. The disease severity classes are defined as follows: 1) Asymptomatic/Paucisymptomatic, 2) Severe, 3) Critical/life-threatening[37].We used this information to investigate if we could identify any contribution of the singleton burden of the prioritized genes to the classification. A multinomial analysis with R was performed using age, gender and the singleton count as explanatory variables. We also analyzed the contribution of the prioritized genes to the outcome (Survived/Deceased) with a logistic regression model and the same covariates used in the multinomial analysis. We performed the analyses using both the whole-gene singleton count and the coding regions singleton count. A summary of the phenotype information is available in Table S4.

## Results

### Population stratification

As expected, the PCA analysis (Figure 1) showed a clear differentiation between QGP and INGI (European) ancestry. The Italian cohorts clustered with the European samples from the 1000Genomes Project reference data, while the QGP samples overlapped with clusters from different populations (AFR, SAS, AMR, EUR). Using the ancestry inference function provided by the KING software, we confirmed the presence of different sub-population clusters in the QGP cohort, highlighting that a considerable proportion of the analyzed samples (more than 4000 samples) belong to a ‘missing’ super population cluster (Figure S1). This is mainly due to the absence of population from the Near East in the 1000Genomes Project data. This outcome confirms results already obtained by different studies on the Italian populations[36] and on the first subset of nearly 6000 samples of the Qatari population[38].

**Figure 1:**
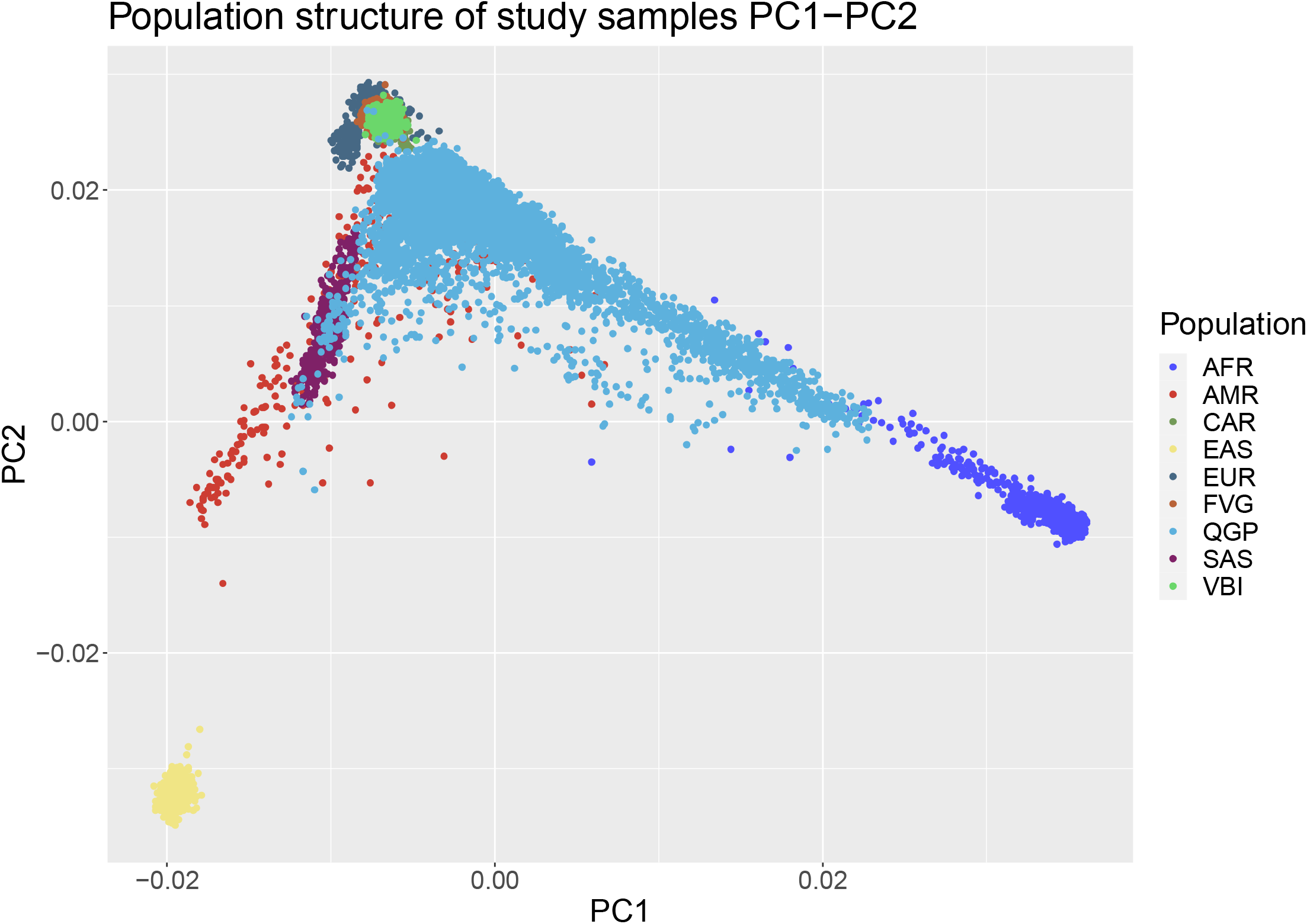
PCA plot of the QGP and INGI cohorts projected onto 1000Genomes Project data. As expected, the first two principal components already show the separation between the QGP and the INGI cohorts and the overlap with the selected populations for the ancestry-related comparisons.

### Population based gene prioritization

For each population and each gene in the selected subset, we calculated two scores related to the presence of cohort-singletons variants. In Figure 2, we show the distribution, among the 3617 genes selected, of the DSC score (Figure 2 top panel) and the SSC score (Figure 2 bottom panel), in each study cohort and the selected reference groups (EUR, AFR and SAS). We compared each target population score distribution with the relevant reference population (Table S5). All the INGI populations are significantly different from the reference EUR population for both scores. In contrast, the Qatari population significantly differs from the reference populations (AFR and SAS) in DSC score distribution, but not for the SSC score distribution. This pattern is also confirmed in the enrichment analyses of relaxed and constrained genes, in target populations versus reference populations, of relaxed and constrained genes. Exact Fisher’s tests show enrichment in constrained genes in all the target populations when comparing DSC scores (Table S6). Similarly, if we consider the SSC scores, all target populations do not show significant enrichment in constrained genes. Regarding the relaxed genes, though, we detected a significant enrichment in both DSC and SSC scores for the Italian populations (CAR, VBI and FVG) but not for the QGP cohort (Table S6). We used a threshold of −2 to define significant constraint and a threshold of +2 to define a significant relaxation signal[39]. Results for the comparisons between our target populations are summarized in Table 1 and Table 2. Regarding the DSC score, we identified six genes with a signature of constraint in the QGP population and relaxation in at least one of the Italian populations. Two of those genes (*TTN* and *LRP1B*) are results of population-specific comparisons, and one (RICTOR) is the outcome of an ancestry-related comparison. Eight genes showed an opposite pattern of relaxation in the Qatari cohort and constraint in at least one Italian cohort. Among them, *RYR3* is the result of an ancestry related comparison. When comparing our target populations based on the overall burden of singletons in each gene (SSC score), we identified a total of 35 genes that behave differently between the Qatari population and at least one Italian population (Table 2). Seventeen of those genes showed a pattern of constraint in the Qatar population and relaxation in at least one Italian cohort. The *HELZ* gene was the only one arising from a population-specific comparison. The remaining eighteen genes showed a pattern of relaxation in the QGP dataset but a significant constraint in at least one of the other targets. In this subset, the CELSR2 gene is the result of a population-specific comparison. Since our focus is to identify genes that consistently show different selection signals among our cohorts, we selected a subset of genes for which both DSC and SSC scores are concordant: *ABCA7, FLNA, MUC5AC* (Table 3). Those three genes showed a consistent relaxation pattern in the QGP cohort while being always characterized by strong signals of constraints in at least one Italian cohort. Interestingly, *FLNA* shows a significant signal of constraint in CAR and VBI cohorts while remaining neutral in the FVG dataset. The trend for the constraint signal is also replicated in all the reference cohorts selected. Data from other outbred populations from the 1000Genome project (EAS and AMR) confirm the trend of constraints (Table S7). *ABCA7* repeats the pattern observed for *FLNA*, in terms of target populations, with a significant constraint signal in the FVG cohort, and a trend of constraint in the CAR cohort, while being neutral in the VBI cohort. This time though, we can see how the outbred reference populations, plus the remaining super populations of 1000Genomes, are all in agreement, showing relaxation signals. Lastly, the *MUC5AC* gene shows a consistent pattern of significant constraint signal in all the Italian cohorts, but conversely, always a significantly relaxed pattern in all other populations.

**Figure 2:**
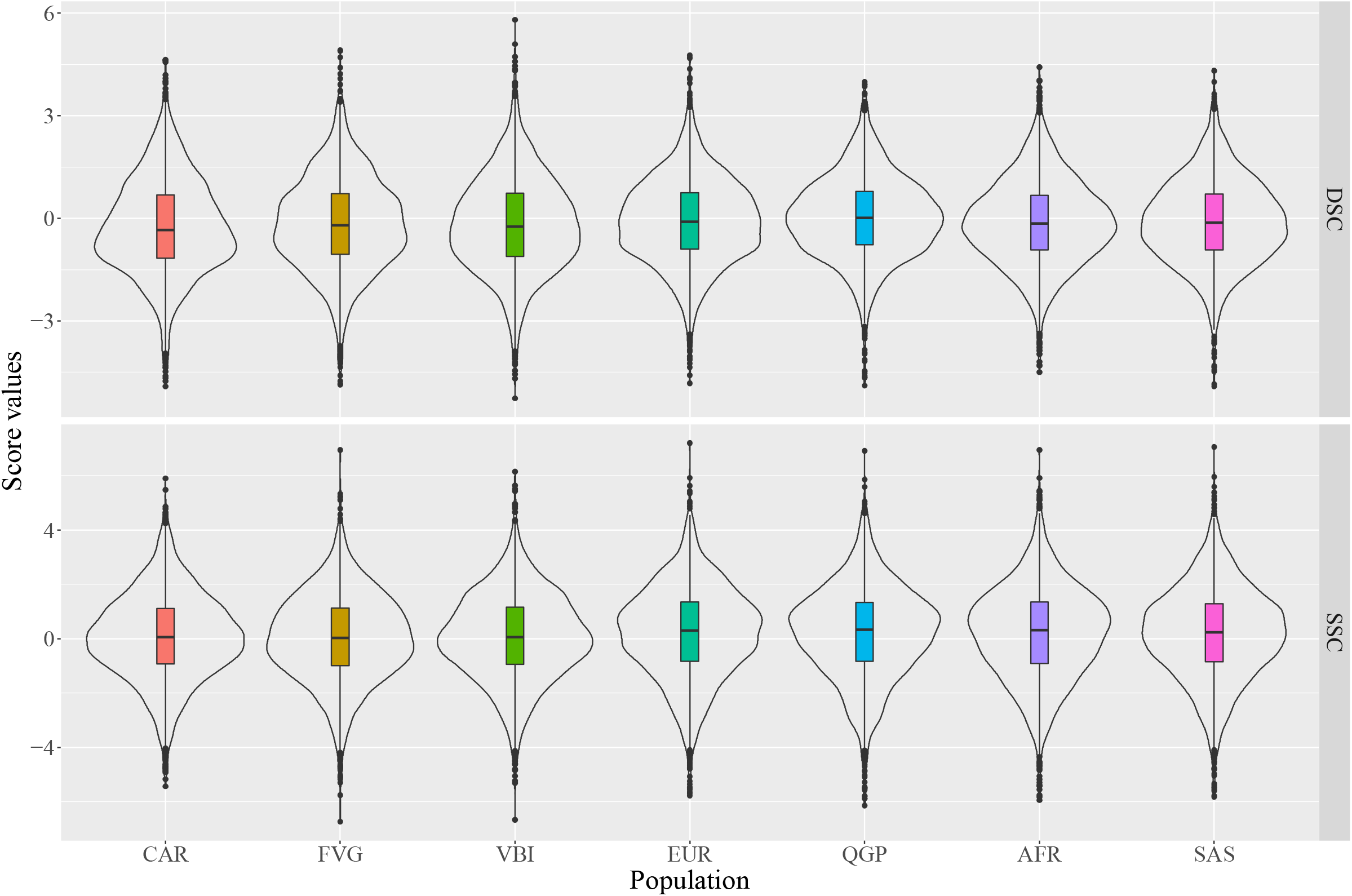
Distributions of the prioritization scores. Violin plots of the distributions of DSC (top panel) and SSC (bottom panel) scores in the subset of selected genes for all target populations (CAR, FVG, VBI, QGP) and all reference outbred populations (AFR, EUR, SAS) from 1000Genomes project.

**Table 1:** Results from comparison of DSC scores between target cohorts (CAR, FVG, VBI, QGP) and the relevant reference superpopulations from the 1000 Genomes Project (EUR, AFR, SAS). The last column refers to the nature of the comparison carried out, as detailed in Supplementary table 3.

| Transcript id | Gene name | DSC score |  |  |  |  |  |  | Comparison |
| --- | --- | --- | --- | --- | --- | --- | --- | --- | --- |
|  |  | QGP | CAR | FVG | VBI | EUR | AFR | SAS |  |
| ENST00000369850 | *FLNA* | 3.854 | -2.435 | 0.272 | -2.510 | -2.399 | -2.166 | -1.879 | C5 |
| ENST00000350763 | *TNC* | 3.370 | -3.792 | 1.388 | -0.631 | 2.666 | 1.838 | 2.187 | C4 |
| ENST00000389048 | *ALK* | 2.575 | 3.651 | 0.290 | -4.098 | 3.388 | 3.212 | 2.852 | C4 |
| ENST00000263094 | *ABCA7* | 2.566 | -0.433 | -2.168 | 0.071 | 3.020 | 2.681 | 2.150 | C4 |
| ENST00000647814 | *ABCC2* | 2.528 | -3.466 | 0.467 | 3.004 | 2.562 | 2.877 | 2.508 | C4 |
| ENST00000621226 | *MUC5AC* | 2.435 | -2.404 | -2.017 | -2.892 | 3.477 | 3.500 | 3.032 | C4 |
| ENST00000634891 | *RYR3* | 2.229 | -3.377 | -1.554 | -3.586 | -2.431 | 2.639 | -3.449 | C8 |
| ENST00000542267 | *FBXL17* | 2.026 | -1.232 | 0.180 | -2.658 | 3.086 | 0.266 | 2.477 | C4 |
| ENST00000589042 | *TTN* | -2.242 | -2.595 | 3.411 | 4.584 | -2.388 | -1.965 | 3.498 | C1 |
| ENST00000357387 | *RICTOR* | -2.369 | -2.206 | -0.033 | 2.407 | 2.181 | 1.284 | -4.070 | C7 |
| ENST00000561890 | *MUC22* | -2.472 | -1.682 | -1.632 | 2.156 | -2.562 | -2.191 | -2.425 | C3 |
| ENST00000336596 | *EPHA3* | -3.001 | 2.139 | -3.421 | -1.535 | -3.704 | 1.921 | -2.814 | C3 |
| ENST00000648947 | *INO80* | -3.444 | -1.309 | 2.424 | -2.928 | -3.439 | -2.692 | -0.727 | C3 |
| ENST00000389484 | *LRP1B* | -4.888 | -4.916 | 3.192 | 2.602 | -4.245 | -2.136 | 3.231 | C1 |

**Table 2:**
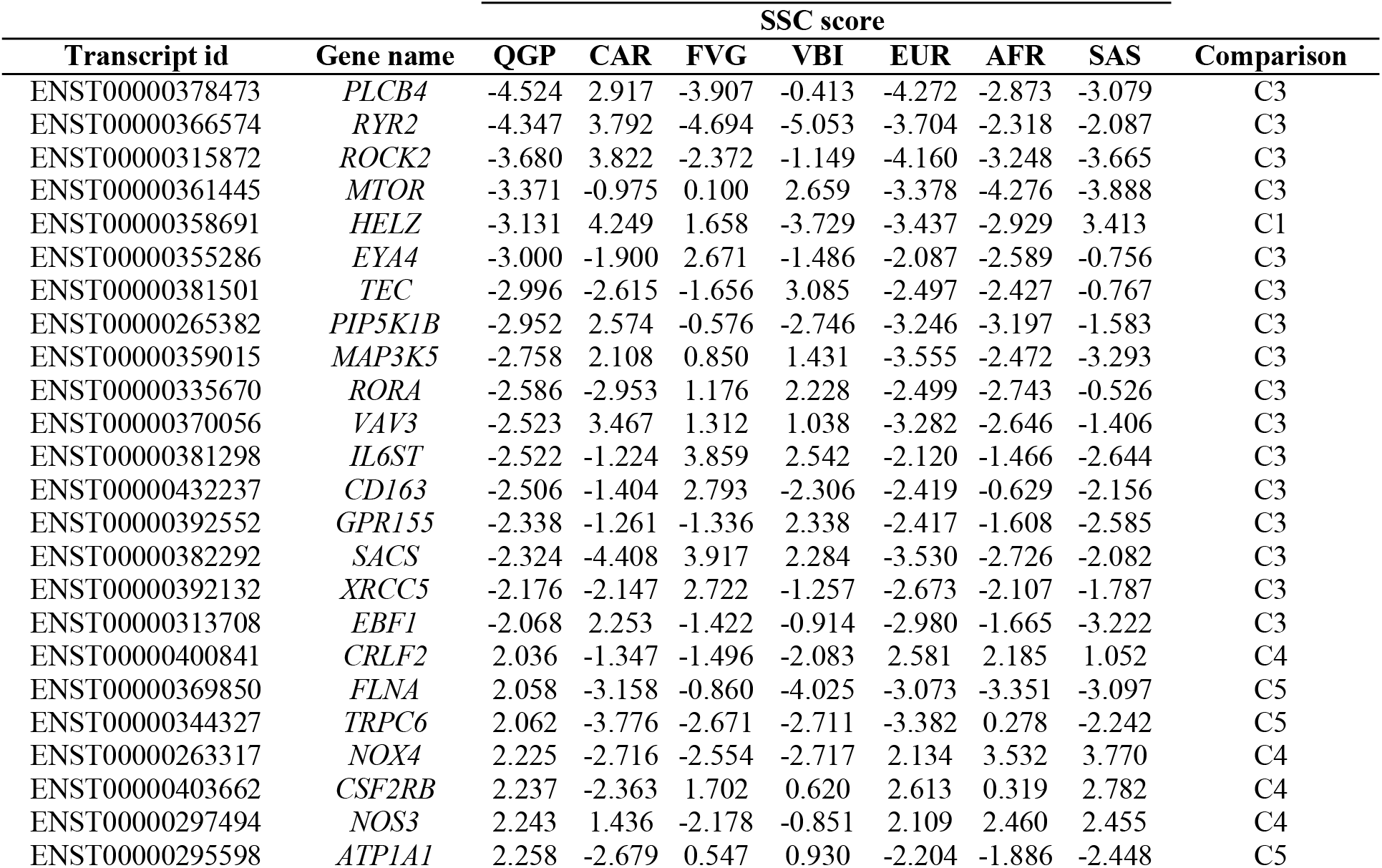
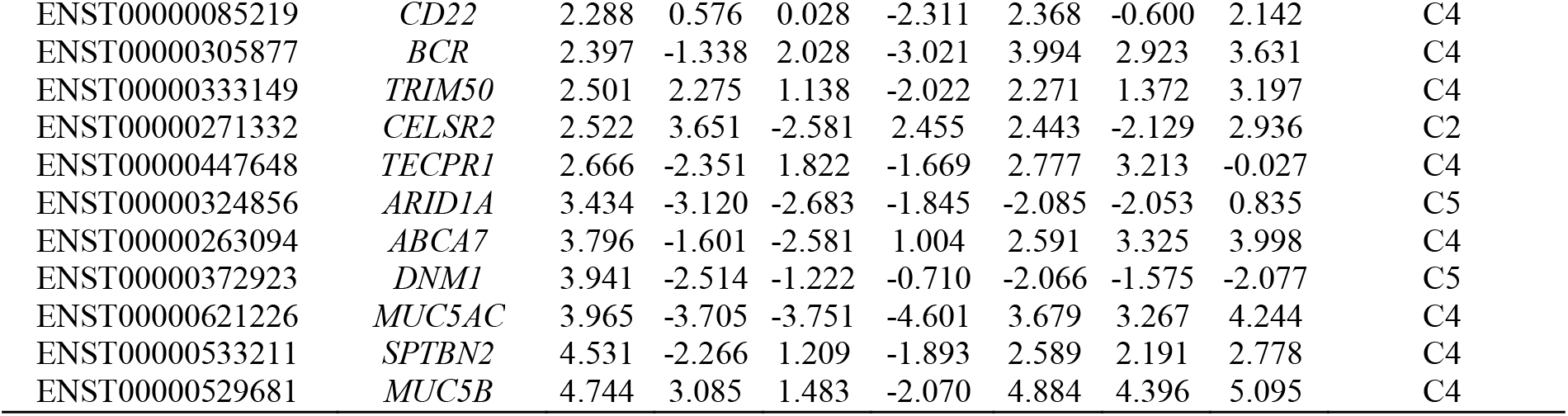
Results from comparison of SSC scores between target cohorts (CAR, FVG, VBI, QGP) and the relevant reference superpopulations from the 1000 Genomes Project (EUR, AFR, SAS). The last column refers to the nature of the comparison carried out, as detailed in Supplementary table 3.

**Table 3:**
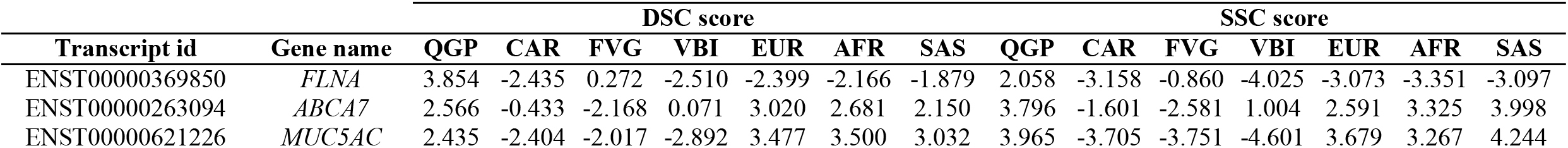
List of genes with a concordant signature of selection between DSC and SSC scores, after the comparison between target cohorts (CAR, FVG, VBI, QGP) and the relevant reference superpopulations from the 1000 Genomes Project (EUR, AFR, SAS).

### Covid-19 cohort analysis

Next, we included a cohort of 189 COVID-19 positive samples (TOV cohort) and calculated the number of singleton variants in this subset for the three genes of interest. Table 4 shows the results of the comparisons with the other study populations and the reference populations. If we consider the whole gene, we can see how, for the *FLNA* gene, the TOV cohort shows a small difference in the singleton density when compared to the FVG cohort and a more significant difference with the VBI and QGP cohorts, while *ABCA7* and *MUC5AC* genes have consistently a significantly different pattern when compared with all reference and target populations (Table S8). If we consider only the coding part of each gene, we confirm the minor differences in the *FLNA* gene between the TOV cohort and the VBI and QGP cohorts. We also confirm the results for *ABCA7* and *MUC5AC* (Table S9).

**Table 4:** Comparison of Singleton burden between the Covid-19 positive cohort (TOV) and other target and reference populations. The reported p-values refer to the comparison between whole gene singleton burden (“P-value whole gene” column) and coding regions singletons burden (“P-value CDS region” column). All singleton counts have been adjusted considering the sample size of each cohort.

| Transcript ID | Gene name | Cohort | P-value whole gene | P-value CDS regions |
| --- | --- | --- | --- | --- |
| ENST00000369850 | FLNA | CAR | 0.630140 | 0.409653 |
|  |  | FVG | 0.046901 | 0.316565 |
|  |  | VBI | 0.000458 | 0.013015 |
|  |  | QGP | 0.000028 | 0.039803 |
|  |  | EUR | 0.312323 | 0.786342 |
|  |  | AFR | 0.878006 | 0.787767 |
|  |  | SAS | 0.200408 | 0.813561 |
| ENST00000263094 | ABCA7 | CAR | 3.2746E-11 | 2.5959E-07 |
|  |  | FVG | 3.1278E-23 | 1.0535E-17 |
|  |  | VBI | 7.1607E-21 | 2.0060E-16 |
|  |  | QGP | 6.2413E-63 | 1.5606E-40 |
|  |  | EUR | 4.4467E-10 | 1.0966E-08 |
|  |  | AFR | 1.7360E-08 | 5.3713E-09 |
|  |  | SAS | 2.3435E-04 | 3.7924E-06 |
| ENST00000621226 | MUC5AC | CAR | 7.4274E-12 | 8.4692E-11 |
|  |  | FVG | 1.8836E-31 | 2.7623E-24 |
|  |  | VBI | 2.3148E-36 | 3.2118E-30 |
|  |  | QGP | 1.5512E-76 | 1.2101E-64 |
|  |  | EUR | 3.3142E-06 | 4.0071E-08 |
|  |  | AFR | 2.9701E-08 | 1.0241E-10 |
|  |  | SAS | 7.2316E-03 | 9.9471E-05 |

To investigate if the burden of singletons in the prioritized genes could contribute to the disease severity classification, we performed multinomial logistic regression analyses. Disease severity class was the response variable, and age, gender and singleton count the explanatory variables. When we consider the contribution of the burden of singleton in the whole gene, the multinomial analyses showed that only age and gender are important predictors for the disease severity classification (Table S10, Figure S2). Using the burden of singletons in the coding regions as parameters in the regression model resulted in age and gender being significant predictors for being in class 2 versus class 1 (p-values: 6.09E-07 and 0.04463 respectively) and belonging to class 3 versus class 1 (p-values: 6.42E-10 and 0.02035). Age resulted in a significant predictor of being in class 2 versus class 3 (p-value: 0.008617). Regarding genes contribution, the model highlighted only the *FLNA* gene as a significant predictor for being in class 2 versus class 1 and for being in class 3 versus class 2 (all p-values: ¡ 2.2e-16) (Table S11).

We also retrieved the disease outcome (Survived/Deceased) information and used the same parameters to perform a logistic regression analysis. As a result, we can still see that age is a major predictor for the outcome (p-value: 1.50e-10) (Figure S3). We can also see a contribution of the *ABCA7* gene, but only when we consider the number of singletons in the whole gene (p-value: 0.0228) (Table S12 and Table S13).

## Discussion

Since the H1N1 influenza pandemic in 1918, the ongoing COVID-19 pandemic is the most severe emergency we have met globally.

For the scientific community, this emergency has been a wake-up call to join forces to fight back, investigate the effect of the virus on patients’ health, and understand the infection’s molecular mechanisms. All this ongoing effort is producing knowledge that is driving therapy and vaccine development. In this context, we focused on population-based statistics to characterize a subset of genes involved in the inflammation/immune response biological process. These statistics were obtained by analyzing populations with different ancestries and levels of inbreeding and consanguinity.

We performed a comparison between our study cohorts and their matched reference populations, according to principal component analysis (PCA) results. The comparisons within our target populations allowed us to show the different patterns in genetic constraints for a large subset of genes involved in the immune response, leading to the prioritization of a group of genes that we could define as “the most differentiated” in terms of signatures of genetic constraints. These differences are most prominent for three genes (*MUC5AC, ABCA7, FLNA*), which harbor a pattern of relaxation in the QGP cohort with respect to other cohorts analyzed. This different pattern of relaxation could be a hint for a different impact of the role of these genes in different populations. Two of the genes, *MUC5AC* and *FLNA*, have already been linked to the COVID-19 host response to different degrees. The *MUC5AC* gene is a gelforming mucin expressed in the lungs in response to infectious agents. This protein plays a protective role against inhaled pathogens, like influenza[40]. A recent study[41] compared levels of MUC5AC, MUC1 and MUC1-CT between critical ill COVID-19 patients and healthy controls, finding a significantly higher level of those proteins in the patients’ mucus. It is also worth noting that another recent work from Kousathanas et al. reported a significant genome-wide association between variants in the MUC1 gene and critical illness caused by[41, 42].

The second gene, *FLNA*, codes for the Filamin A protein, which has been identified as a putative interaction candidate with coronaviruses S protein and is involved in the coronavirus replication cycle[43]. A recent study showed that the *FLNA* gene is part of the host protein-protein interaction (PPI) network for the SARS-CoV-2 virus and among the targets of different drugs under development[44]. A loss-of-function mutation of the *FLNA* gene was reported in family adults with emphysema[45]. There is no study showing a direct link between the I gene and COVID-19 yet, but it has been proven that it is highly expressed in the reticuloendothelial system and modulates the phagocytosis activity[46, 47], though its function, like many other ABC-transporters, has yet to be clarified. Interestingly the BioGRID interactome database[48] lists physical interactions of *ABCA7* with *ADBR2, C5AR2* and *SGTB*, among others. Each one of these genes has been linked to COVID-19 host response in previous studies, in terms of interaction[49], therapy[50] and severity in case of pre-existing health conditions[51].

Altogether with the information available on the prioritized genes and the knowledge of the different evolution of the pandemic between Qatar and Italy, we performed a proof-of-concept analysis. Using the information provided by a cohort of COVID-19 positive samples from Italy, to try to identify, if present, the contribution of the amount of ultra-rare variants in those genes to the outcome of the disease (Survived/Deceased) and the severity (Asymptomatic/Paucisymptomatic, Severe, Critical/life-threatening).

From a cohort-based perspective, we can see differences in the distribution of singletons in the COVID-19 positive samples regarding our study populations and their reference populations. This outcome suggests that those three genes could play a role in the description of the cohort and that investigating rare genetic variations occurring in those genetic regions could be a starting point to complete the characterization of those samples. With a subsequent approach, we applied logistic regression analyses to investigate the impact of the singleton burden in the three prioritized genes on the disease outcome and disease severity. While the contribution of age and sex is explicit and expected, these analyses suggested that the burden of singletons carried by each patient in the *ABCA7* gene could predict a worse outcome together with age. From the point of view of the disease severity, the burden of singleton in the *FLNA* gene could help discriminate samples with distinct levels of disease severity. In this case, though, our findings seem to be inconsistent since we find that having a lower burden of singletons is a predictor of developing a severe reaction. However, a high singleton burden is a predictor of developing a critical reaction. This finding can be better explained by looking at the distribution of singletons in our cohort stratified by disease severity. For the *FLNA* gene, all samples belonging to class 2 of severity do not carry any singleton. This feature could be introduced by one of the limitations of this study: the sample size of the COVID-19 positive cohort. Increasing the number of cases will undoubtedly allow us to have a better estimate of the singleton distribution. Moreover, in our model, we didn’t include any of the risk factors that are already linked to a diverse response to the infection.

One last limitation could be represented by the inclusion of only one cohort of COVID-19 positive samples, for which only Whole Exome sequence data was available. We chose to include this cohort due to the phenotypical characterization, which allowed us to investigate our hypothesis of a genetic contribution to the disease severity prioritized genes. Nevertheless, for all the cohorts involved, information on the COVID-19 affected samples is already being collected. That will allow us to produce more precise results with further analyses. To our knowledge, this is the first study performing a whole-genome population-level comparison between Arabian and European populations, both differently affected by the pandemic. Recent similar studies focused only on the ACE2 receptor and populations from the 1000Genomes Project[52] or compared allele frequencies on covid-19 related genes in the Brazilian population with data from the 1000Genome and gnomAD datasets[53].

With the development of new vaccines against SARS-CoV-2 infection, we are bound to see a decrease in adverse disease outcomes and disease severity among the immunized populations. However, our work could be a starting point to better prioritize genes that could be therapeutical targets in different populations. Moreover, with the increased knowledge obtained thanks to the many studies that focused on understanding virus-host interaction, we could extend our method to any new similar threat that should arise in the future.

## Conclusion

we were able to identify three candidate genes that could be further investigated for their role in the COVID-19 infection, and we want to stress the message that harnessing the information provided by rare genetic variants, in this still evolving context, is proving increasingly useful to explain the different outcomes of this disease.

## Data Availability

Access to the QGP data used for this study is through a dedicated portal by QGP (Accession ID: QF-QGP-RES-PUB-0226). The informed consent given by the study participants does not cover posting of participant level phenotype and genotype data of Qatar Biobank (QBB)/Qatar Genome Project (QGP) in public databases. Access to QBB/QGP data can be obtained through an established ISO-certified process by submitting a project request at https://www.qatarbiobank.org.qa/research/ how-apply which is subject to approval by the QBB IRB committee.
The genetic data belonging to INGI cohorts and analyzed in this manuscript have been submitted to the European Variation Archive (EVA) are accessible in Variant Call Format (VCF) at the following link: https://www.ebi.ac.uk/ena/data/view/PRJEB33648

## Supplementary information

**Figure S1: Inferred ancestry**. Using the king software inferred ancestry function, we were able to correctly infer the ancestry of our target populations, matching the already available data.

**Figure S2: Disease severity classification in the TOV cohort**. The box plots show that age is still a major discriminant factor related to disease severity, in the TOV cohort. Disease severity classes are defined as 1) Asymptomatic/Paucisymptomatic, 2) Severe, 3) Critical/life-threatening

**Figure S3: Disease outcome classification in the TOV cohort**. Thebox plots show the clear contribution of age, in the definition of the disease outcome classes.

**Table S1:** Initial candidate gene list of 3617 genes curated according to the knowledge-literature base by the Ingenuity® Variant Analysis™ software (https://www.qiagenbioinformatics.com/products/ingenuity-variant-analysis) from QIAGEN, Inc. (IVA) and the viral gene panel expert from Genomics England (GEL).

**Table S2:** List of genes recently associated with COVID-19 susceptibility and severity

**Table S3:** Details of the comparisons defined between Target and Refer ence populations

**Table S4:** Summary statistics of the phenotype information available for the TOV cohort.

**Table S5:** Comparison between scores distribution of target populations versus reference populations

**Table S6:** Enrichment tests in target populations versus reference populations

**Table S7:** DSC and SSC scores for FLNA, MUC5AC and ABCA7 in all 1000Genomes super populations

**Table S8:** Comparison of Gene Singleton Density between the Covid-19 positive cohort (TOV) and other target and reference populations. All singleton counts have been adjusted considering the sample size of each cohort.

**Table S9:** Comparison of Coding Singleton Density between the Covid-19 positive cohort (TOV) and other target and reference populations. All singleton counts have been adjusted considering the sample size of each cohort.

**Table S10:** Multinomial analysis results, comparing classes of disease severity, considering the burden of singletons in the whole gene. Class 1: Asymptomatic/Paucisymptomatic; Class 2: Severe; Class 3: Critical/life-threatening. Only significant predictors are reported.

**Table S11:** Multinomial analysis results, comparing classes of disease severity, considering the burden of singletons in the coding regions of each gene. Class 1: Asymptomatic/Paucisymptomatic; Class 2: Severe; Class 3: Critical/life-threatening. Only significant predictors are reported.

**Table S12:** Logistic analysis results for disease outcome classification.

Only significant predictors are reported.

**Table S13:** Logistic analysis results for disease outcome classification considering the burden of singletons in the coding regions of each gene. Only significant predictors are reported.

## Author Contributions

Conceptualization, M.C. and H.M.; Methodology, M.C., M.M.; Formal Analysis, M.C, Y.AS and C.S., H.M; Resources, D.C., A.N., I.Q., G.N., W.A., A.AT., S.S., A.A., S.I.I.; Writing Original Draft Preparation, H.M., C.M.,Y.AS.,C.S., M.M.; Writing Review Editing, H.M.,C.M.,Y.AS.,C.S., M.M.

## Institutional Review

The recruitment and sequencing of participants from the Qatar Biobank (QBB) were approved by the Hamad Medical Corporation Ethics Committee in 2011 and continued with QBB IRB from 2017 onwards and renewed annually (protocol IRB-A-QBB-2019-0017). The genetic analyses were approved by the QBB IRB protocol E-2020-QBB-Res-ACC-0226-0130.

## Data Availability

Access to the QGP data used for this study is through a dedicated portal by QGP (Accession ID: QF-QGP-RES-PUB-0226). The informed consent given by the study participants does not cover posting of participant level phenotype and genotype data of Qatar Biobank (QBB)/Qatar Genome Project (QGP) in public databases. Access to QBB/QGP data can be obtained through an established ISO-certified process by submitting a project request at https://www.qatarbiobank.org.qa/research/ how-apply which is subject to approval by the QBB IRB committee.

The genetic data belonging to INGI cohorts and analyzed in this manuscript have been submitted to the European Variation Archive (EVA) are accessible in Variant Call Format (VCF) at the following link: https://www.ebi.ac.uk/ena/data/view/PRJEB33648

## Acknowledgments

We would like to thank the Qatar Biobank for data collection and phenotyping. The Qatar Genome Program and Qatar Biobank are both Research, Development Innovation entities within Qatar Foundation for Education, Science and Community Development. We would like to thank the inhabitants and local administrations of Friuli Venezia Giulia Region, Carlantino and Val Borbera that made these studies possible.

