## supplementary table for "Poking COVID-19: insights on genomic constraints among immune-related genes between Qatari and Italian populations"

**Table S2:** List of genes recently associated with COVID-19 susceptibility and severity

| **Gene** | **Variants** | **Source** | **Publication** |
| --- | --- | --- | --- |
| ***SLC6A2*** | rs35081325 | HGI | not published in a peer review journal |
| ***CXCR6*** |  | HGI |  |
| ***CCR1*** |  | HGI |  |
| ***OAS1*** | rs4766664 | HGI |  |
| ***OAS2*** |  | HGI |  |
| ***OAS3*** |  | HGI |  |
| ***DPP9*** | rs2277732 | HGI |  |
| ***TYK2*** | rs7956615 | HGI |  |
| ***IFNAR2*** | rs13050728 | HGI |  |
| ***IL10RB*** |  | HGI |  |
| ***IFNAR1*** |  | HGI |  |
| ***IFNAR2*** |  | HGI |  |
| ***FOXP4*** | rs1853837 | HGI |  |
| ***TLR3*** |  | Human Genetic Effort (HGE) | Zhnag et al, Science 2020 |
| ***UNC93B1*** |  | Human Genetic Effort (HGE) |  |
| ***TICAM1*** |  | Human Genetic Effort (HGE) |  |
| ***TBK1*** |  | Human Genetic Effort (HGE) |  |
| ***IRF3*** |  | Human Genetic Effort (HGE) |  |
| ***IRF7*** |  | Human Genetic Effort (HGE) |  |
| ***LZTFL1*** |  | Genomicc UK | Castineira et al MedriXiv |
| ***CCHCR1*** |  | Genomicc UK |  |
| ***HLA-DPB1*** |  | Genomicc UK |  |
| ***VSTM2A*** |  | Genomicc UK |  |
| ***HAS2-AS1*** |  | Genomicc UK |  |
| ***IGF1*** |  | Genomicc UK |  |
| ***ABO*** |  |  |  |

**Table S3:** Details of the comparisons defined between Target and Reference populations

|  |  | **Target 1** | **Reference 1** |  | **Target 2** | **Reference 2** |
| --- | --- | --- | --- | --- | --- | --- |
| **Population specific** | **C1** | Relaxed (+) | Costrained (-) | vs | Costrained (-) | Relaxed (+) |
|  | **C2** | Costrained (-) | Relaxed (+) | vs | Relaxed (+) | Costrained (-) |
| **Population specific vs ancestry related** | **C3** | Relaxed (+) | Costrained (-) | vs | Costrained (-) | Costrained (-) |
|  | **C4** | Costrained (-) | Relaxed (+) | vs | Relaxed (+) | Relaxed (+) |
|  | **C5** | Costrained (-) | Costrained (-) | vs | Relaxed (+) | Costrained (-) |
|  | **C6** | Relaxed (+) | Relaxed (+) | vs | Costrained (-) | Relaxed (+) |
| **Ancestry related** | **C7** | Relaxed (+) | Relaxed (+) | vs | Costrained (-) | Costrained (-) |
|  | **C8** | Costrained (-) | Costrained (-) | vs | Relaxed (+) | Relaxed (+) |

**Table S 4:** Summary statistics of the phenotype information availabe for the TOV cohort.

|  |  | **Gender** | | **Outcome** | |  |
| --- | --- | --- | --- | --- | --- | --- |
| **Disease Severity group** | **n** | **M (n)** | **F (n)** | **Survived (n)** | **Deceased (n)** | **Age (mean)** |
| Asymptomatic/Paucisymptomatic (1) | 52 | 25 | 27 | 52 | 0 | 37.5 |
| Severe (2) | 73 | 45 | 28 | 60 | 13 | 60.4 |
| Critical/life-threatening (3) | 64 | 40 | 24 | 21 | 43 | 68.4 |
| **total** | 189 | 110 | 79 | 133 | 56 | 57.73 |

**Table S5:** Comparison between scores distribution of target populations versus reference populations

| **Reference** | **Target** | **P-value DSC** | **P-value SSC** |
| --- | --- | --- | --- |
| **EUR** | **CAR** | 1.33E-06 | 7.79E-07 |
| **EUR** | **FVG** | 3.17E-02 | 3.65E-08 |
| **EUR** | **VBI** | 3.78E-03 | 3.84E-07 |
| **AFR** | **QGP** | 4.64E-07 | 4.36E-01 |
| **SAS** | **QGP** | 2.97E-05 | 5.35E-01 |

**Table S6:** Enrichment tests in target populations versus reference populations

|  |  | **DSC score** | | **SSC score** | |
| --- | --- | --- | --- | --- | --- |
| **Reference** | **Target** | **Costrained**  **p-value** | **Relaxed**  **p-value** | **Costrained**  **p-value** | **Relaxed**  **p-value** |
| **EUR** | **CAR** | 2.08E-06 | 6.81E-04 | 1.38E-01 | 7.11E-04 |
| **EUR** | **FVG** | 2.27E-03 | 7.70E-02 | 3.19E-01 | 7.80E-06 |
| **EUR** | **VBI** | 2.27E-03 | 1.93E-03 | 2.82E-01 | 3.68E-06 |
| **AFR** | **QGP** | 3.72E-03 | 4.17E-01 | 1.00E+00 | 8.60E-01 |
| **SAS** | **QGP** | 3.51E-02 | 2.52E-01 | 7.51E-01 | 8.60E-01 |

**Table S7:** DSC and SSC scores for *FLNA*, *MUC5AC* and *ABCA7* in all 1000Genomes superpopulations

| **Gene Name** | **Score** | **AFR** | **AMR** | **EAS** | **EUR** | **SAS** |
| --- | --- | --- | --- | --- | --- | --- |
| ***ABCA7*** | **DSC** | 2.6813 | 1.6151 | 2.5922 | 3.0203 | 2.1504 |
|  | **SSC** | 3.3248 | 4.0146 | 3.7889 | 2.5907 | 3.9976 |
| ***FLNA*** | **DSC** | -2.1657 | -2.0498 | -2.2511 | -2.3994 | -1.8791 |
|  | **SSC** | -3.3509 | -3.3736 | -3.2086 | -3.0729 | -3.0966 |
| ***MUC5AC*** | **DSC** | 3.4995 | 2.8133 | 2.8882 | 3.4774 | 3.0323 |
|  | **SSC** | 3.2673 | 4.0639 | 4.1692 | 3.6789 | 4.2443 |

**Table S8:** Comparison of Gene Singleton Density between the Covid-19 positive cohort (TOV) and other target and reference populations. All singleton counts have been adjusted considering the sample size of each cohort.

| **Transcript ID** | **Gene name** | **Cohort** | **P-value** | **OR** | **TOV Gene singleton count** | **TOV Gene singleton density** | **Cohort Gene singleton count** | **Cohort Gene singleton density** |
| --- | --- | --- | --- | --- | --- | --- | --- | --- |
| **ENST00000369850** | ***FLNA*** | **CAR** | 0.630140 | 1.4201 | 13 | 0.00050 | 6 | 0.00023 |
|  |  | **FVG** | 0.046901 | 2.3537 | 13 | 0.00050 | 11 | 0.00042 |
|  |  | **VBI** | 0.000458 | 5.8145 | 13 | 0.00050 | 5 | 0.00019 |
|  |  | **QGP** | 0.000028 | 4.3237 | 13 | 0.00050 | 229 | 0.00877 |
|  |  | **EUR** | 0.312323 | 0.6893 | 13 | 0.00050 | 61 | 0.00234 |
|  |  | **AFR** | 0.878006 | 0.9218 | 13 | 0.00050 | 60 | 0.00230 |
|  |  | **SAS** | 0.200408 | 0.6464 | 13 | 0.00050 | 61 | 0.00234 |
| **ENST00000263094** | ***ABCA7*** | **CAR** | 3.2746E-11 | 10.9752 | 84 | 0.00328 | 5 | 0.00020 |
|  |  | **FVG** | 3.1278E-23 | 12.8380 | 84 | 0.00328 | 13 | 0.00051 |
|  |  | **VBI** | 7.1607E-21 | 8.5370 | 84 | 0.00328 | 22 | 0.00086 |
|  |  | **QGP** | 6.2413E-63 | 17.8135 | 84 | 0.00328 | 359 | 0.01404 |
|  |  | **EUR** | 4.4467E-10 | 2.9802 | 84 | 0.00328 | 91 | 0.00356 |
|  |  | **AFR** | 1.7360E-08 | 2.5141 | 84 | 0.00328 | 142 | 0.00555 |
|  |  | **SAS** | 2.3435E-04 | 1.8442 | 84 | 0.00328 | 138 | 0.00540 |
| **ENST00000621226** | ***MUC5AC*** | **CAR** | 7.4274E-12 | 7.4508 | 114 | 0.00264 | 10 | 0.00023 |
|  |  | **FVG** | 1.8836E-31 | 15.0962 | 114 | 0.00264 | 15 | 0.00035 |
|  |  | **VBI** | 2.3148E-36 | 19.6004 | 114 | 0.00264 | 13 | 0.00030 |
|  |  | **QGP** | 1.5512E-76 | 15.4443 | 114 | 0.00264 | 562 | 0.01301 |
|  |  | **EUR** | 3.3142E-06 | 1.9908 | 114 | 0.00264 | 185 | 0.00428 |
|  |  | **AFR** | 2.9701E-08 | 2.2129 | 114 | 0.00264 | 219 | 0.00507 |
|  |  | **SAS** | 7.2316E-03 | 1.4702 | 114 | 0.00264 | 235 | 0.00544 |

**Table S9:** Comparison of Coding Singleton Density between the Covid-19 positive cohort (TOV) and other target and reference populations. All singleton counts have been adjusted considering the sample size of each cohort.

| **Transcript ID** | **Gene name** | **Cohort** | **P-value** | **OR** | **TOV CDS singleton count** | **TOV CDS singleton density** | **Cohort CDS singleton count** | **Cohort CDS singleton density** |
| --- | --- | --- | --- | --- | --- | --- | --- | --- |
| **ENST00000369850** | ***FLNA*** | **CAR** | 0.409653 | 3.2705 | 5 | 0.00063 | 1 | 0.00013 |
|  |  | **FVG** | 0.316565 | 1.9922 | 5 | 0.00063 | 5 | 0.00063 |
|  |  | **VBI** | 0.013015 | 11.1693 | 5 | 0.00063 | 1 | 0.00013 |
|  |  | **QGP** | 0.039803 | 2.7799 | 5 | 0.00063 | 137 | 0.01725 |
|  |  | **EUR** | 0.786342 | 1.1543 | 5 | 0.00063 | 14 | 0.00176 |
|  |  | **AFR** | 0.787767 | 1.1814 | 5 | 0.00063 | 18 | 0.00227 |
|  |  | **SAS** | 0.813561 | 0.7582 | 5 | 0.00063 | 20 | 0.00252 |
| **ENST00000263094** | ***ABCA7*** | **CAR** | 2.5959E-07 | 14.3697 | 44 | 0.00683 | 2 | 0.00031 |
|  |  | **FVG** | 1.0535E-17 | 43.6709 | 44 | 0.00683 | 2 | 0.00031 |
|  |  | **VBI** | 2.0060E-16 | 19.6584 | 44 | 0.00683 | 5 | 0.00078 |
|  |  | **QGP** | 1.5606E-40 | 24.9877 | 44 | 0.00683 | 134 | 0.02080 |
|  |  | **EUR** | 1.0966E-08 | 4.0565 | 44 | 0.00683 | 35 | 0.00543 |
|  |  | **AFR** | 5.3713E-09 | 3.8931 | 44 | 0.00683 | 48 | 0.00745 |
|  |  | **SAS** | 3.7924E-06 | 2.9601 | 44 | 0.00683 | 45 | 0.00699 |
| **ENST00000621226** | ***MUC5AC*** | **CAR** | 8.4692E-11 | 12.4070 | 76 | 0.00448 | 4 | 0.00024 |
|  |  | **FVG** | 2.7623E-24 | 18.8715 | 76 | 0.00448 | 8 | 0.00047 |
|  |  | **VBI** | 3.2118E-30 | 42.4223 | 76 | 0.00448 | 4 | 0.00024 |
|  |  | **QGP** | 1.2101E-64 | 23.1499 | 76 | 0.00448 | 250 | 0.01474 |
|  |  | **EUR** | 4.0071E-08 | 2.6675 | 76 | 0.00448 | 92 | 0.00542 |
|  |  | **AFR** | 1.0241E-10 | 3.1351 | 76 | 0.00448 | 103 | 0.00607 |
|  |  | **SAS** | 9.9471E-05 | 1.9678 | 76 | 0.00448 | 117 | 0.00690 |

**Table S10:** Multinomial analysis results, comparing classes of disease severity, considering the burden of singletons in the whole gene. Class 1: Asymptomatic/Paucisymptomatic; Class 2: Severe; Class 3: Critical/life-threatening. Only significant predictors are reported.

|  |  |  |  |  |  | **IC 95%** | |  |
| --- | --- | --- | --- | --- | --- | --- | --- | --- |
| **Severity Class** | **Predictor** | **Estimate** | **Std. Error** | **P-value** | **OR** | **Lower bound** | **Upper bound** | **Std. Error (OR)** |
| **Class 2 vs Class 1** | **Age** | 6.72E-02 | 1.36E-02 | 8.35E-07 | 1.0695029 | 1.0412933 | 1.0984768 | 0.0145859 |
|  | **Gender (Male)** | 9.25E-01 | 4.63E-01 | 0.05 | 2.5208194 | 1.0173269 | 6.2463014 | 1.1670436 |
| **Class 3 vs Class 1** | **Age** | 1.01E-01 | 1.63E-02 | 4.92E-10 | 1.1064869 | 1.0717711 | 1.1423270 | 0.0179959 |
|  | **Gender (Male)** | 1.10E+00 | 5.09E-01 | 0.03 | 2.9987634 | 1.1061065 | 8.1299424 | 1.5259327 |
| **Class 3 vs Class 2** | **Age** | 3.40E-02 | 1.17E-02 | 0 | 1.0345794 | 1.0111462 | 1.0585557 | 0.0120932 |

**Table S11:** Multinomial analysis results, comparing classes of disease severity, considering the burden of singletons in the coding regions of each gene. Class 1: Asymptomatic/Paucisymptomatic; Class 2: Severe; Class 3: Critical/life-threatening. Only significant predictors are reported.

|  |  |  |  |  |  | **IC 95%** | |  |
| --- | --- | --- | --- | --- | --- | --- | --- | --- |
| **Severity Class** | **Predictor** | **Estimate** | **Std. Error** | **P-value** | **OR** | **Lower bound** | **Upper bound** | **Std. Error (OR)** |
| **Class 2 vs Class 1** | **Age** | 6.87E-02 | 1.38E-02 | 6.09E-07 | 1.0711545 | 1.0426111 | 1.1004793 | 0.0147605 |
|  | **Gender (Male)** | 9.39E-01 | 4.68E-01 | 0.04 | 2.5573716 | 1.0227587 | 6.3946161 | 1.1958014 |
|  | ***FLNA*** | -1.04E+01 | 7.39E-05 | < 2.2e-16 | 0.0000296 | 0.0000296 | 0.0000296 | 0.0000000022 |
| **Class 3 vs Class 1** | **Age** | 9.86E-02 | 1.60E-02 | 6.42E-10 | 1.1036457 | 1.0696605 | 1.1387107 | 0.0176120 |
|  | **Gender (Male)** | 1.18E+00 | 5.08E-01 | 0.02 | 3.2520969 | 1.2007352 | 8.8080493 | 1.6532035 |
| **Class 3 vs Class 2** | **Age** | 2.99E-02 | 1.14E-02 | 0.01 | 1.0303340 | 1.0076149 | 1.0535653 | 0.0117211 |
|  | ***FLNA*** | 7.98E+00 | 5.18E-01 | < 2.2e-16 | 2933.6216596 | 1063.3832398 | 8093.1650220 | 1518.8708798 |

**Table S12:** Logistic analysis results for disease outcome classification. Only significant predictors are reported.

|  |  |  |  |  |  | **IC 95%** | |  |
| --- | --- | --- | --- | --- | --- | --- | --- | --- |
| **Outcome** | **Predictor** | **Estimate** | **Std. Error** | **P-value** | **OR** | **Lower bound** | **Upper bound** | **Std. Error (OR)** |
| **Survied vs Deceased** | **Age** | 1.07E-01 | 1.67E-02 | 1.50E-10 | 1.1129343 | 1.0770954 | 1.1499656 | 0.0185860 |
|  | **ABCA7** | 4.20E-01 | 1.85E-01 | 0.0228 | 1.5225705 | 1.0603341 | 2.1863118 | 0.2810665 |

**Table S13:** Logistic analysis results for disease outcome classification considering the burden of singletons in the coding regions of each gene. Only significant predictors are reported.

|  |  |  |  |  |  | **IC 95%** | |  |
| --- | --- | --- | --- | --- | --- | --- | --- | --- |
| **Outcome** | **Predictor** | **Estimate** | **Std. Error** | **P-value** | **OR** | **Lower bound** | **Upper bound** | **Std. Error (OR)** |
| **Survied vs Deceased** | **Age** | 1.05E-01 | 1.66E-02 | 2.33E-10 | 1.1110439 | 1.0754556 | 1.1478098 | 0.0184544 |
